# Time From Witnessed Cardiac Arrest to First Defibrillation and Population-Level Implications of Earlier Defibrillation: A Nationwide Cohort Study in Japan

**DOI:** 10.64898/2026.11.13.26367394

**Authors:** Atsushi Kubo, Shinobu Tamura, Daigo Morioka, Kenko Fukui, Atsushi Hiraide

## Abstract

**Background:** Early defibrillation is the principal modifiable determinant of survival after shockable out-of-hospital cardiac arrest, but studies restricted to defibrillated patients cannot observe those who had degenerated before a responder arrived. We quantified this loss and its population-level consequences.

**Methods:** We analysed 368,496 bystander-witnessed adult arrests of presumed cardiac origin in the All-Japan Utstein Registry (2005–2024). Modified Poisson models with marginal standardization estimated the proportion with a shockable rhythm at emergency medical services (EMS) contact and 1-month survival as functions of elapsed time. Assuming exchangeability within strata of bystander category and age, we estimated the additional survivors had patients reached at ≥15 minutes been reached at 5–10 minutes.

**Results:** The adjusted proportion with a shockable rhythm was 33.1% (95% CI, 30.2–36.5) at 1 minute, 24.7% (24.4–24.9) at 9 minutes and 8.4% (8.0–8.8) at 30 minutes, a relative decline of 4.8% per minute between 5 and 30 minutes. Adjusted 1-month survival fell from 39.5% (38.4– 40.6) at 9 minutes to 15.9% (15.1–16.7) at 20 minutes after EMS defibrillation, and from 63.5% (59.9–67.3) at 3 minutes to 47.2% (44.4–50.0) at 10 minutes after bystander defibrillation; each minute cost 2.1 to 2.7 percentage points in both pathways. Shifting the 119,982 patients reached at ≥15 minutes to 5–10 minutes yielded 6,465 additional survivors (6,259–6,669) over 20 years (323 per year; number needed to treat, 18.6), 59.1% through rhythm preservation. Family-witnessed arrests and the “other” bystander category, predominantly institutional, accounted for 77.4%.

**Conclusions:** The proportion of patients shockable at EMS contact was approximately 5% lower for each additional minute after collapse, and survival fell by about two percentage points per minute whoever delivered the shock. Under the model’s assumptions, most of the benefit of faster contact derives from reaching patients while still shockable, chiefly in arrests witnessed by family members or in institutional settings.

## Introduction

Early defibrillation is the single most time-critical intervention in the management of out-of-hospital cardiac arrest (OHCA) caused by a shockable rhythm.^1^ Nationwide dissemination of public-access automated external defibrillators in Japan has been accompanied by an increase in the proportion of patients receiving a bystander shock and by improved neurologically favorable survival at the population level.^2,3^ The underlying rationale is that survival after ventricular fibrillation declines rapidly from the moment of collapse, and that shortening the interval to the first shock is therefore the principal modifiable determinant of outcome.^4,5,6^

Most evidence supporting this rationale, however, has been generated from studies defined by the receipt of defibrillation. Nationwide Japanese analyses have compared recipients and non-recipients of public-access defibrillation and, more recently, the timing of public-access defibrillation among patients with ventricular fibrillation in public locations.^3,7^ A recent analysis of time-synchronized defibrillator recordings found that the proportion of ventricular fibrillation episodes terminated by the first shock fell from 93% to 75% as the delay to that shock lengthened, and concluded that the mechanisms underlying the survival penalty of delay have not yet been elucidated.^8^ Such designs condition on a patient having been in a shockable rhythm when a responder arrived. They are therefore well suited to estimating how much is lost by delivering a shock later to a patient who remains shockable, but they cannot observe the patients who were shockable at the moment of collapse and had already degenerated to asystole or pulseless electrical activity by the time anyone was in a position to act. If the interval to first contact is long, this second group may be large, and the benefit of a faster system may lie substantially in preventing patients from leaving the population eligible for defibrillation at all.

Two questions therefore remain unresolved. First, how rapidly does the probability of remaining in a shockable rhythm decline after a witnessed collapse, and how does this compare with the decline in survival among those who are defibrillated. Second, if the interval from collapse to first responder contact were shortened at a population level, how much of the resulting benefit would arise from each of these two mechanisms, and how large would the total be.

Using a nationwide, population-based registry covering 20 years of OHCA in Japan, we estimated the rate at which shockable rhythms are lost with elapsed time, characterized the relationship between the interval to first defibrillation and survival, and translated these observed relationships into population-level counterfactual scenarios in which first contact occurred earlier than observed.

## Methods

### Design and data source

We conducted a nationwide, population-based observational cohort study using the All-Japan Utstein Registry, a prospective registry of OHCA maintained by the Fire and Disaster Management Agency of Japan, covering all arrests attended by emergency medical services (EMS) nationwide according to standardized Utstein-style definitions.^9^ Records from 1 January 2005 through 31 December 2024 were analyzed. Exact duplicate records (n=210) were removed, and 18 records filed in the 2005 registry file whose year field was coded as 2004, all with event dates in 2005, were assigned to 2005. The study follows the STROBE reporting guideline.^10^

### Study population

We identified witnessed arrests of presumed cardiac origin in adults aged 18 years or older. The registry classifies bystanders as family members, friends, colleagues, passers-by or other. Two further exclusions were applied on the basis of the recorded event sequence.

First, we excluded arrests witnessed by fire department or EMS personnel, in whom EMS contact preceded or coincided with the recorded witness time in 96.8%, because the time structure of resuscitation differs fundamentally (Supplementary Methods S2).

Second, we excluded patients in whom the emergency call preceded the witnessed collapse, indicating that EMS had already been activated for another reason; in these patients the interval from collapse to contact is mechanically shortened (Supplementary Methods S3).

All eligible patients were included; no sample size calculation was performed.

### Timestamp quality control

Timestamps were assessed independently, without assuming which of two inconsistent timestamps was erroneous. Witness times were classified by calendar consistency against the EMS contact time, and defibrillation recorded after hospital arrival was treated as logically inconsistent. Problematic timestamps were set to missing, with one exception: because every analysis depends on the time of collapse, patients were excluded (Supplementary Methods S1).

### Exposure definitions

The bystander automated external defibrillator fields changed in 2021. Before 2021, bystander defibrillation was a binary indicator whose accompanying timestamp did not record the shock: the timestamp’s prevalence tracked that of bystander chest compression to within 1.2 percentage points between 2005 and 2012. From 2021 a four-level field was accompanied by a timestamp of shock delivery. Across the change the proportion receiving bystander defibrillation was continuous, but the recorded interval from collapse shifted 3 to 5 minutes later at every quantile with no change in survival, so the bystander-pathway analysis was restricted to 2021–2024. The EMS defibrillation timestamp was stable throughout (median 14 minutes in both periods) and was analyzed for the full period (Supplementary Methods S4).

For the rhythm-decay analysis the exposure was the interval from collapse to EMS contact, when the rhythm is first documented, among patients reached within 40 minutes who had not received a bystander shock and in whom ventricular fibrillation, pulseless ventricular tachycardia, pulseless electrical activity or asystole was documented. The population-level analysis used the same interval and does not depend on either defibrillation timestamp.

### Outcomes

The primary outcome was 1-month survival; the secondary outcome was a favorable neurological outcome (Cerebral Performance Category 1 or 2) at 1 month.

### Analysis A: observed timing–outcome association

The two delivery pathways occupied almost non-overlapping exposure ranges and served different populations, so separate models were fitted over each pathway’s range of support: 0–15 minutes for bystander defibrillation (2021–2024) and 9–40 minutes for EMS defibrillation (2005–2024). The EMS lower bound of 9 minutes was chosen because, after exclusion of call-before-collapse cases, shorter intervals comprised a heavily selected residual (Supplementary Methods S3).

Within each pathway, 1-month survival was modeled by modified Poisson regression with a log link,^11^ with elapsed time modeled as a restricted cubic spline and, for the bystander pathway, a separate indicator for shocks recorded at zero minutes. Models were adjusted for age (spline, 4 df), sex, bystander category, bystander chest compression (performed versus not recorded as performed), and, for the EMS pathway, calendar year (spline, 4 df), indicators for 2013 onward and 2021 onward, when recorded bystander intervention variables changed, an indicator for the pandemic period 2020–2022, and prefecture (Supplementary Methods S4–S5). Predicted probabilities were obtained by marginal standardization over the observed covariate distribution,^12^ with 95% confidence intervals from 500 nonparametric bootstrap replicates in which model fitting was repeated; covariate rate ratios used robust variance (Supplementary Table S3). Models were fitted in patients with complete covariates (complete-case analysis; Supplementary Methods S6). The rhythm-decay analysis used the same framework, with a shockable rhythm at EMS contact as the outcome and the covariates listed in the Figure 2 legend; bystander chest compression was not included (Supplementary Methods S8). Category-specific curves (Supplementary Figure S1) came from a model that added a bystander category × elapsed-time interaction.

### Analysis B: population-level counterfactual

We estimated the additional 1-month survivors expected if patients reached by EMS at 15 minutes or later had instead been reached at 5–10 minutes. Both intervals lie within the observed data, so no extrapolation was required; each interval excludes its upper bound.

Estimates were computed within joint strata defined by bystander category and age group (<65, 65–74, 75–84, ≥85 years) and summed, because the late-contact group was 2 to 3 years older than the early-contact group within the two largest bystander categories (Supplementary Table S1). For stratum s, with N patients in the late group, p-late and p-early the proportions with a shockable rhythm at contact, s-late and s-early 1-month survival among those with a shockable rhythm, and n-late survival among those without, Channel A was defined as N × p-late × (s-early − s-late) and Channel B as N × (p-early − p-late) × (s-early − n-late).

Channel A represents earlier defibrillation among patients shockable in either scenario; Channel B represents patients who would have been shockable had contact occurred earlier.^13^ Patients non-shockable in both scenarios were assigned no benefit, so the estimate excludes the general benefit of earlier EMS arrival. This is a decomposition of a difference between two rates into a rate component and a composition component,^14^ and the allocation follows from a monotonicity assumption: patients shockable at the later contact time are assumed to have been shockable at the earlier time. The decomposition is exact and leaves no interaction residual (Supplementary Methods S7). Uncertainty was propagated by drawing each component 20,000 times from its binomial sampling distribution; intervals are the 2.5th and 97.5th percentiles, untruncated. The number needed to treat (NNT) was the target population size divided by the estimated additional survivors.

### Sensitivity analyses

Sensitivity analyses comprised a conservative counterfactual target of 10–15 minutes; an upper limit of 120 minutes, or no upper limit, for the interval to EMS contact; restriction of the target population to patients reached at 20 minutes or later; restriction to 2005–2019, 2015–2019 and 2020–2024; retention of call-before-collapse patients; omission of the age stratification; exclusion of patients who received bystander defibrillation; and restriction of the EMS pathway to 2021–2024 to match the bystander period. Results are given below and in Supplementary Table S4 and Supplementary Figure S5.

### Software and ethics

Analyses used R version 4.5.2 with the data.table, splines, sandwich, lmtest and ggplot2 packages. Analytic code is provided in the Supplement. This study was approved by the Institutional Review Board of Meiji University of Integrative Medicine (No. 2026-030). The requirement for individual informed consent was waived by the IRB due to the observational and retrospective nature of the use of anonymized registry data. Registry data usage was authorized by the Fire and Disaster Management Agency (FDMA) of Japan.

## Results

### Study population

Of 2,482,808 records in the registry files for 2005–2024, 210 exact duplicates were removed. Of the remaining 2,482,598 OHCAs, 1,485,558 were of presumed cardiac origin, of which 596,363 were witnessed and 592,508 occurred in adults. We excluded 114,498 arrests witnessed by fire department or EMS personnel and 105,137 patients (22.0%) in whom the emergency call preceded the witnessed collapse, leaving 372,873 bystander-witnessed adult arrests of presumed cardiac origin. After exclusion of 4,377 patients (1.2%) with missing or implausible time records, the analytic population comprised 368,496 (Figure 1).

**Figure 1.**
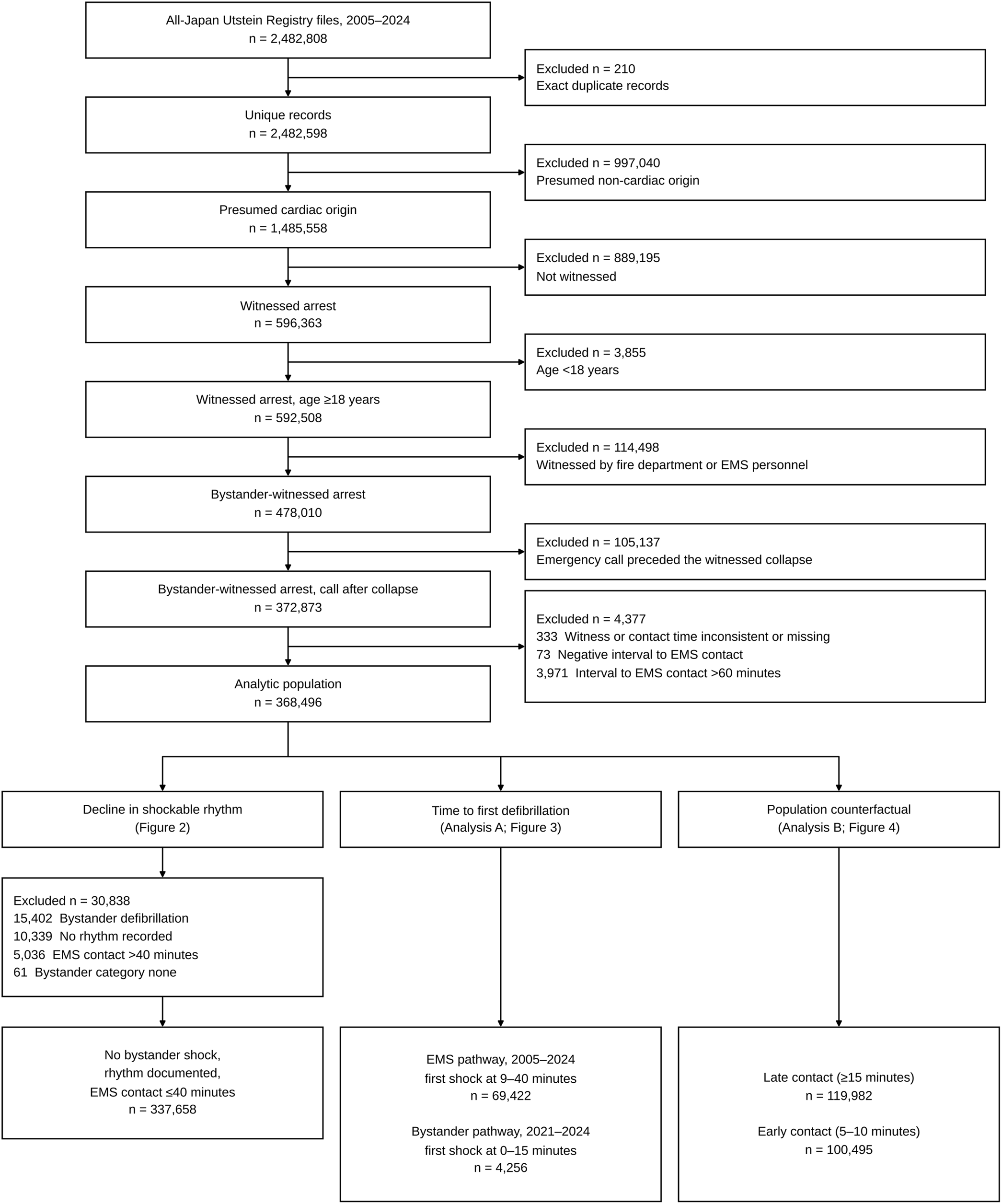
Study flow. EMS denotes emergency medical services. Models in Analysis A included patients with complete covariates; 5 EMS-pathway patients were excluded for missing covariates (Supplementary Methods S6).

Among 151,849 patients with linked ambulance transport records, 92.3% of family-witnessed arrests occurred in a residence and 57.1% of colleague-witnessed arrests at a workplace, and 84.2% of arrests in the “other” category were classed as public premises, 84.7% of which were a hospital, clinic or nursing home (Supplementary Table S6).

### Decline in shockable rhythm with elapsed time

Among 337,658 patients who did not receive bystander defibrillation, in whom a rhythm was documented at EMS contact and who were reached within 40 minutes of collapse, the proportion with a shockable rhythm declined monotonically with elapsed time, from 43.5% within 2 minutes of collapse to 25.9% at 8–10 minutes, 14.2% at 16–20 minutes, and 6.0% at 31–40 minutes (Figure 2). After standardization, the adjusted proportion fell from 33.1% (95% CI, 30.2–36.5) at 1 minute, where data were sparse, to 28.6% (28.0–29.1) at 5 minutes, 24.7% (24.4– 24.9) at 9 minutes, 18.0% (17.7–18.2) at 15 minutes, 13.4% (13.1–13.7) at 20 minutes, and 8.4% (8.0–8.8) at 30 minutes, corresponding to a relative decline of approximately 4.8% per minute between 5 and 30 minutes.

**Figure 2.**
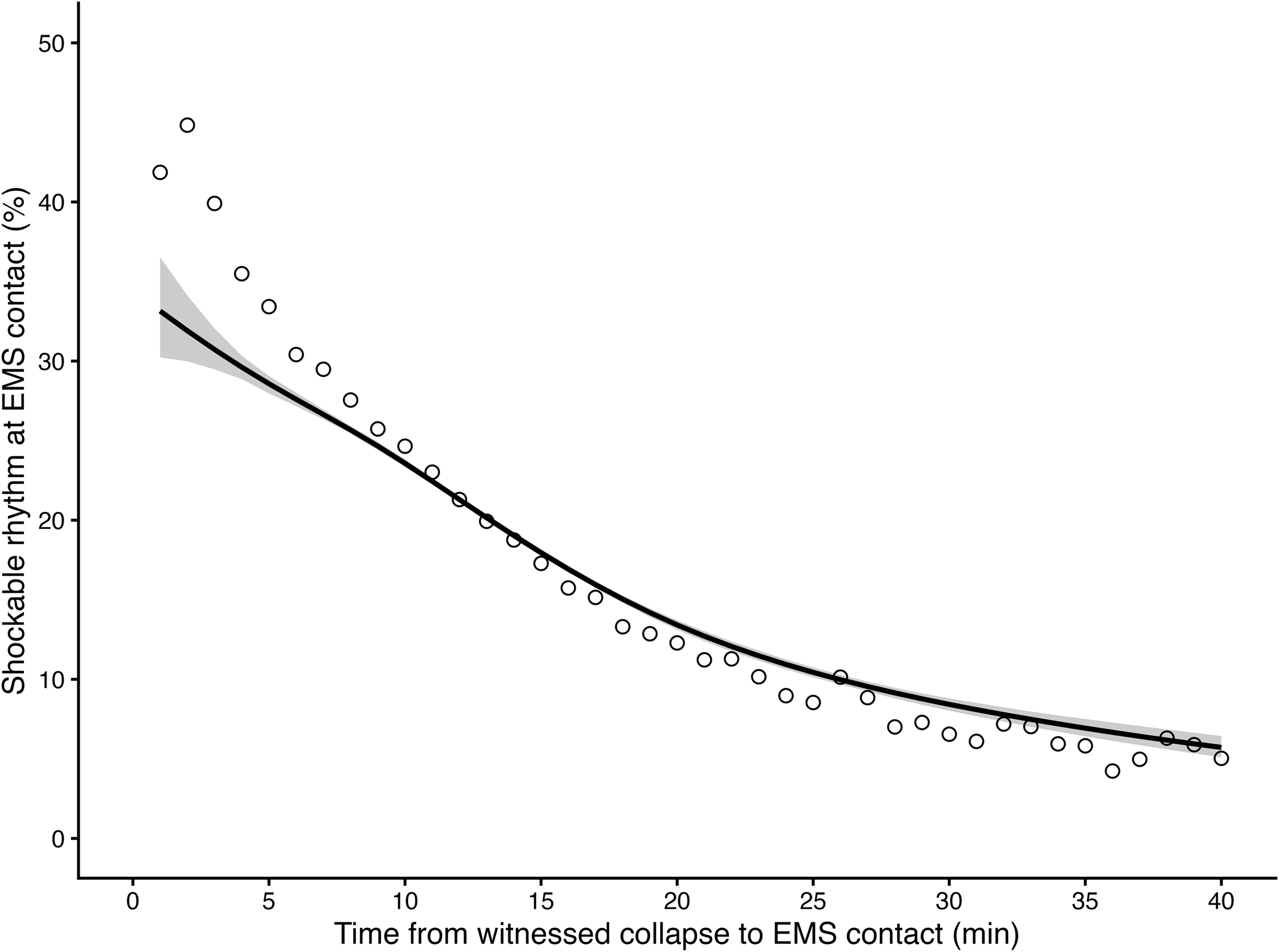
Proportion of patients with a shockable rhythm at emergency medical services contact according to time from witnessed cardiac arrest, among patients who did not receive bystander defibrillation and were reached within 40 minutes, 2005–2024 (n=337,658). The line shows marginally standardized predicted probabilities from a modified Poisson model adjusted for age, sex, bystander category, calendar year, and indicators for 2013 onward, 2021 onward and 2020– 2022; the shaded area is the 95% confidence interval from 500 bootstrap replicates. Open circles are observed proportions for each minute, with 0 and 1 minute combined. Observed values exceed the standardized curve at short intervals because patients reached early were younger.

The rate of decline differed by bystander category (P for interaction <0.001). At 9 minutes, the proportion with a shockable rhythm standardized to age 78 years was 27.9% for colleague-witnessed and 27.4% for friend-witnessed arrests, compared with 20.5% for family-witnessed and 17.9% for the “other” category, which predominantly comprised institutional settings (Supplementary Figure S1).

### Time to first defibrillation and survival

The bystander pathway (2021–2024) comprised 4,270 patients and the EMS pathway (2005– 2024) 76,748. The two pathways served markedly different populations (Table 1): bystander-defibrillated patients were less often witnessed by a family member (7.4% versus 65.2%), more often in the “other” category (43.0% versus 13.4%), and almost universally received bystander chest compressions (98.6% versus 56.3%). Median time to first defibrillation was 5 minutes (IQR 2–8) and 14 minutes (IQR 11–19), respectively. One-month survival was 54.2% and 25.1%, and favorable neurological outcome 45.6% and 15.7%.

**Table 1.** Characteristics of patients receiving first defibrillation, by pathway.

| **Characteristic** | **Bystander pathway**<br>**2021–2024 (n=4,270)** | **EMS pathway**<br>**2005–2024 (n=76,748)** |
|---|---|---|
| Age, years, median (IQR) | 66 (55–78) | 69 (58–78) |
| Male sex | 3,455 (80.9) | 59,841 (78.0) |
| Bystander category |  |  |
| Family member | 315 (7.4) | 50,053 (65.2) |
| Friend | 548 (12.8) | 5,574 (7.3) |
| Colleague | 826 (19.3) | 6,308 (8.2) |
| Passer-by | 743 (17.4) | 4,527 (5.9) |
| Other | 1,838 (43.0) | 10,286 (13.4) |
| Bystander chest compression | 4,210 (98.6) | 43,188 (56.3) |
| Shockable rhythm at EMS contact† | 1,093 (25.6) | 59,967 (78.1) |
| Time to first defibrillation, min, median (IQR) | 5 (2–8) | 14 (11–19) |
| 1-month survival | 2,313 (54.2) | 19,295 (25.1) |
| Cerebral Performance Category 1–2‡ | 1,947 (45.6) | 12,071 (15.7) |

Values are n (%) unless otherwise indicated. IQR denotes interquartile range; EMS, emergency medical services. The bystander pathway is restricted to 2021–2024, when the registry records the time of shock delivery.
† Rhythm recorded at the time of EMS patient contact. In the bystander-defibrillation group this reflects the rhythm after the bystander shock had already been delivered and therefore does not represent the initial rhythm; it is not comparable between pathways.
‡ Includes 54 patients in the EMS pathway recorded with a category of 1 or 2 despite documented death at one month, retained as recorded (Supplementary Methods S6). Among survivors, a category of 1 or 2 was recorded in 84.2% of the bystander pathway and 62.3% of the EMS pathway.

Adjusted, marginally standardized survival declined steeply with elapsed time in both pathways (Figure 3). In the bystander pathway, survival was highest within the first 4 minutes (63.5% at 3 minutes; 95% CI, 59.8–67.7) and fell to 57.3% (54.7–60.0) at 5 minutes, 47.0% (44.2–49.8) at 10 minutes and 34.3% (28.5–39.4) at 14 minutes, a relative decline of 5.5% per minute between 3 and 14 minutes. Survival at 0 minutes was 49.9% (44.7–55.3), lower than at 1–4 minutes.

**Figure 3.**
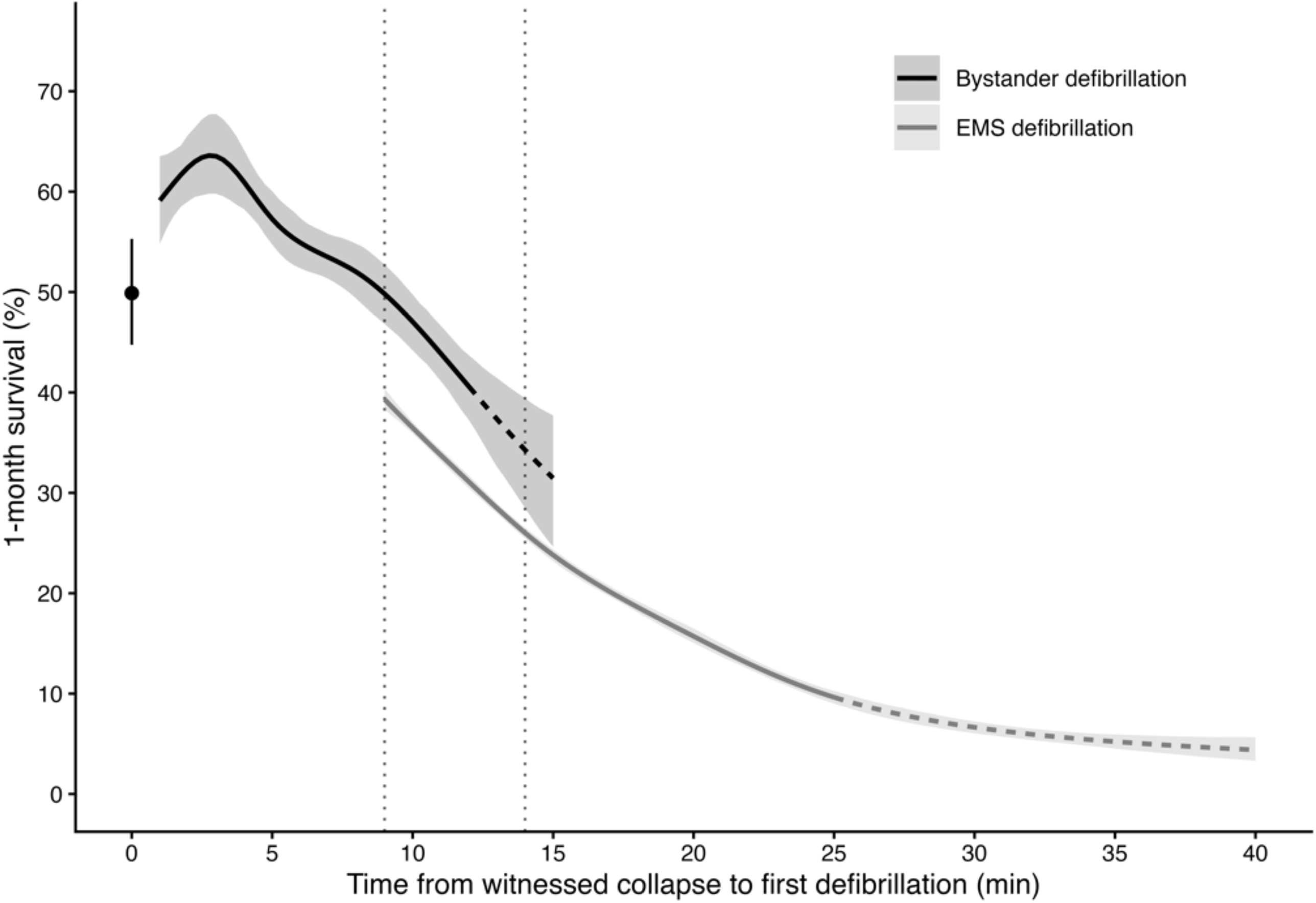
Adjusted 1-month survival according to time from witnessed cardiac arrest to first defibrillation, by defibrillation pathway (bystander pathway, 2021–2024, n=4,270; emergency medical services pathway, 2005–2024, n=76,748). Curves are marginally standardized predicted probabilities from modified Poisson models; shaded areas are 95% confidence intervals from 500 nonparametric bootstrap replicates. Dotted vertical lines delimit the interval in which both pathways have data (9–14 minutes). Curves are dashed where data are sparse or estimates unstable: beyond 12 minutes for the bystander pathway and beyond 25 minutes for the emergency medical services pathway. The point at 0 minutes, with its 95% confidence interval, is shown separately because shocks recorded at zero minutes were modeled with a separate indicator.

In the EMS pathway, survival fell from 39.3% (38.3–40.4) at 9 minutes to 31.1% (30.5–31.7) at 12 minutes, 23.8% (23.2–24.4) at 15 minutes, 15.7% (15.1–16.5) at 20 minutes and 6.6% (6.0– 7.2) at 30 minutes, a relative decline of 8.0% per minute between 9 and 20 minutes.

Where both pathways had data (9–14 minutes), survival after bystander defibrillation exceeded survival after EMS defibrillation by 8.3 to 10.6 percentage points, with non-overlapping confidence intervals at every minute (49.8% versus 39.3% at 9 minutes; 47.0% versus 36.5% at 10 minutes; 40.6% versus 31.1% at 12 minutes; 34.3% versus 26.0% at 14 minutes). Restricting the EMS analysis to 2021–2024 to match the bystander period narrowed the difference from 10.8 percentage points at 9 minutes to 6.8 at 14 minutes, with non-overlapping confidence intervals through 13 minutes.

### Population-level implications of earlier defibrillation

Under the primary counterfactual scenario, in which the 119,982 patients reached by EMS at 15 minutes or later had instead been reached at 5–10 minutes, we estimated 6,465 additional 1-month survivors (95% CI, 6,259–6,669) over the 20-year period, or 323 per year, corresponding to an NNT of 18.6 (Figure 4).

**Figure 4.**
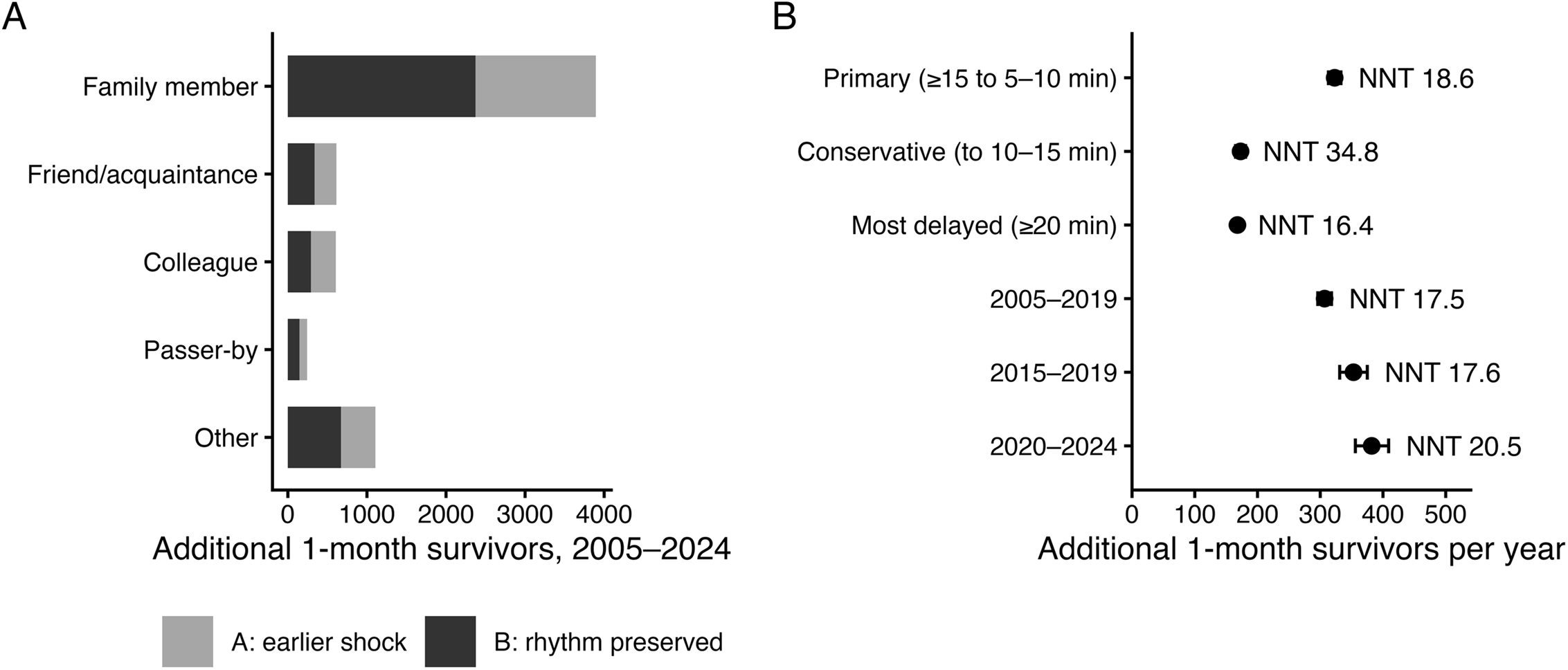
Estimated population-level impact of earlier first emergency medical services contact. (A) Additional 1-month survivors over the 20-year study period under the primary counterfactual scenario (n=119,982 patients reached at 15 minutes or later), decomposed by bystander category and by mechanism. Channel A represents earlier defibrillation among patients already in a shockable rhythm; Channel B represents patients who would still have been in a shockable rhythm had contact occurred earlier. The “other” category predominantly comprised institutional settings. (B) Annualized estimates across counterfactual scenarios, with 95% intervals from Monte Carlo propagation of binomial sampling distributions. NNT denotes the number of patients whose contact interval would need to be shortened to yield one additional survivor.

The rhythm-preservation channel accounted for 3,821 additional survivors (3,690–3,953, 59.1%), compared with 2,644 (2,511–2,776, 40.9%) from earlier defibrillation among patients already in a shockable rhythm. Rhythm preservation accounted for the majority of the benefit in every scenario examined (54.3% to 67.7%; Supplementary Table S4). Family-witnessed arrests accounted for 3,895 additional survivors (60.2% of the total) and the “other” category for 1,107 (17.1%), together representing 77.4% of the estimated benefit.

Repeating the analysis with a favorable neurological outcome as the endpoint yielded 4,835 additional patients with Cerebral Performance Category 1 or 2 (4,661–5,009; 242 per year; NNT, 24.8), equivalent to 74.8% of the additional survivors, of which rhythm preservation accounted for 55.9%.

### Sensitivity analyses

The direction of findings was consistent across scenarios (Figure 4B and Supplementary Table S4). A more conservative target of 10–15 minutes yielded 3,452 additional survivors (3,291– 3,612; 173 per year; NNT 34.8). Restricting the target population to the 55,028 patients reached at 20 minutes or later yielded 3,361 (3,265–3,459; 168 per year) with the most favorable efficiency (16.4) and the highest contribution from rhythm preservation (67.7%). Omitting the age stratification raised the primary estimate to 7,890 (7,657–8,122; 15.2). Excluding patients who received bystander defibrillation, in whom the rhythm at EMS contact is a post-treatment variable, gave 6,368 (6,173–6,566; 18.1), and retaining patients in whom the call preceded the collapse gave 5,280 (5,107–5,457; 23.2).

## Discussion

In this nationwide study of 368,496 bystander-witnessed adult arrests over 20 years, the proportion of patients shockable at EMS contact was approximately 4.8% lower in relative terms for each additional minute after collapse, and survival after defibrillation fell by 2.1 to 2.7 percentage points per minute whoever delivered the shock. Shifting the 119,982 patients reached at 15 minutes or later to 5–10 minutes would have yielded an estimated 6,465 additional 1-month survivors, or 323 per year (NNT, 18.6), three-fifths of them through reaching patients while they were still shockable.

### Loss of defibrillation eligibility as the dominant mechanism

The decomposition of the counterfactual into two channels is the principal contribution of this analysis. Conventional framing of early defibrillation emphasizes the time-dependent decline in the success of a shock once delivered. Our data indicate that this mechanism accounts for approximately two-fifths of the potential population benefit. The remainder reflects patients who, by the time the first responder arrived, had already degenerated from a shockable to a non-shockable rhythm and therefore never became candidates for defibrillation at all. These patients are invisible to any study restricted to defibrillation recipients.^3,7^ This allocation rests on a monotonicity assumption, that patients in a shockable rhythm when reached late would also have been shockable if reached earlier; an alternative algebraic partition of the same total would assign 23.2% rather than 59.1% to rhythm preservation (Supplementary Methods S7). The total estimate does not depend on this choice.

The time-dependent fall in first-shock success that constitutes Channel A has been demonstrated directly with defibrillator recordings, each additional minute of delay being associated with a 6% higher probability of failure to terminate ventricular fibrillation.^8^ That analysis was restricted to patients presenting with ventricular fibrillation as the initial rhythm, a design that measures Channel A with far greater physiological precision than a registry can but that by construction cannot observe Channel B, because patients who had already left the shockable state before any rhythm was recorded are absent from its population. The component that analysis could not observe is, under the monotonicity assumption, the larger of the two.

The share attributable to rhythm preservation rose as fewer patients remained eligible: it was lowest for colleague-witnessed arrests (48.1%), in which 34.9% of patients remained shockable at late contact compared with 12.3% of family-witnessed arrests, and highest when the target population was restricted to patients reached at 20 minutes or later (67.7%). Because Channel A is proportional to the fraction still shockable at late contact, this gradient partly reflects the structure of the decomposition.

### Relation to previous nationwide Japanese studies

Earlier analyses of this registry linked the dissemination of public-access defibrillation to improved population-level neurological survival without defining an individual time origin,^2,3^ and a recent analysis matched on the timing of public-access defibrillation among recipients with ventricular fibrillation in public locations.^7^ Our analysis adds residential and institutional arrests, quantifies the loss of defibrillation eligibility directly, and extends the observed relationships to a population-level counterfactual (Supplementary Table S2).

### Two pathways, two populations

At the same elapsed time, survival after bystander defibrillation was 8 to 11 percentage points higher than after EMS defibrillation (7 to 11 percentage points within the common period 2021– 2024), a difference consistent with earlier comparisons of defibrillation by bystanders and EMS.^15,16^ It cannot be attributed to the act of defibrillation itself: bystander-defibrillated patients were less often at home and almost all received chest compressions. The two pathways therefore identify different populations, and a pooled time-response curve would be uninterpretable.

### Where the remaining burden lies

Although the per-patient benefit of earlier defibrillation was greatest in workplace and public settings, 77.4% of the estimated population benefit arose from family-witnessed arrests (60.2%) and arrests in the “other” category (17.1%), because these settings account for most cases.

Current public-access defibrillation policy in Japan has concentrated devices in transport hubs, schools and commercial premises, consistent with randomized and observational evidence of benefit in public locations.^17,18^ Our findings indicate that the largest remaining opportunity lies in the settings that these bystander categories predominantly represent, residences and institutions, where response intervals are longest and the loss of defibrillation eligibility before first contact is correspondingly greatest; dispatch of nearby lay responders is one means of shortening this interval.^19,20^ The annual size of the target population grew from 5,385 (2005–2019 average) to 6,212 (2015–2019) and 7,843 (2020–2024; Supplementary Figure S4), and the expected annual benefit from 307 to 353 and 382 additional survivors, while the NNT rose modestly from 17.5 to 20.5. The absolute burden is therefore increasing with population aging, and the growth is concentrated in the groups in which the per-patient opportunity is smallest. These observed estimates complement a previous projection based on synthetic patient data.^21^

### Concentration of pre-activated patients in short-interval strata

In 22.0% of bystander-witnessed arrests the emergency call preceded the witnessed collapse. Although this proportion was stable at 21–23%, these patients, whose interval to contact is mechanically shortened, became concentrated in short-interval strata as contact intervals lengthened, accounting for 41.0% of the 5–10 minute stratum in 2022 compared with 23.1% in 2005 (Supplementary Figure S2). Because they were deteriorating under medical attention and less often shockable, including them flattens the apparent survival curve at short intervals (Supplementary Figure S3). Analyses using the witnessed collapse as time zero, including those relating bystander interventions to ambulance response time,^22^ should identify and handle this group explicitly.

### Limitations

The registry did not record the location of arrest, and bystander category served as a proxy. Linkage of ambulance transport records for 2016–2019 and 2021–2022 confirmed that this proxy identifies residences and institutional settings with high fidelity (Supplementary Table S6), but it was not available for the full study period and could not be used for adjustment; residual confounding by location therefore remains possible, particularly in the comparison between pathways. The counterfactual rests on an exchangeability assumption that cannot be verified.^23^ Within bystander category, patients reached late were 2 to 3 years older than those reached early in the two largest categories, and estimates were therefore computed within joint strata of bystander category and age group; omitting this stratification would have raised the primary estimate from 6,465 to 7,890. Bystander cardiopulmonary resuscitation was more frequent in the late group in every category, which biases the estimate conservatively.

The reported confidence intervals reflect sampling variability alone and do not incorporate uncertainty in the exchangeability assumption; the range across scenarios that retained the age stratification, from 168 to 384 additional survivors per year, is a more realistic expression of uncertainty. The bystander-pathway estimates, confined to 2021–2024, are less precise. The rhythm-decay curve is cross-sectional: it compares different patients observed at different intervals rather than tracking individual patients over time.

Among patients defibrillated by EMS, the proportion with a shockable rhythm at EMS contact fell from 94.8% at 9–11 minutes to 27.0% at 26–40 minutes (Supplementary Table S5), indicating that late defibrillations increasingly represent arrhythmia arising during resuscitation rather than delayed treatment of a persistent shockable rhythm; estimates beyond 25 minutes are not interpreted. In the bystander pathway, survival at zero minutes was 14 percentage points below that at 3 minutes. Patients shocked at zero minutes were older than those shocked at 1–4 minutes (median, 70 versus 66 years) and more often in the “other” category (58.0% versus 49.0%), consistent with residual confounding by institutional setting and frailty among patients for whom a device was already in place. Finally, the counterfactual assumes that a 5–10 minute contact interval is attainable for all patients currently reached later, which is not realistic in rural areas; the more conservative 10–15 minute scenario, yielding 173 additional survivors per year, may better represent what is operationally achievable. Generalizability to EMS systems with different response intervals, dispatch practices or termination-of-resuscitation rules is uncertain.

## Conclusions

Among bystander-witnessed cardiac arrests of presumed cardiac origin in Japan, the proportion of patients in a shockable rhythm at EMS contact was approximately 5% lower in relative terms for each additional minute after collapse, and survival after defibrillation fell by roughly two percentage points per minute whoever delivered the shock. Advancing first contact from 15 minutes or more to 5–10 minutes would be expected to produce approximately 320 additional survivors per year nationally, with the majority of that benefit deriving from the preservation of defibrillation eligibility rather than from earlier delivery of a shock to patients already eligible. Judged by bystander category, the largest remaining opportunity lies in residential and institutional settings.

## Non-standard Abbreviations and Acronyms

EMS: emergency medical services
NNT: number needed to treat
OHCA: out-of-hospital cardiac arrest

## Acknowledgments

The authors thank the Fire and Disaster Management Agency of Japan for maintaining the nationwide emergency medical service transport records and making these data available for research.

## Sources of Funding

This research received no specific grant from any funding agency in the public, commercial, or not-for-profit sectors.

## Disclosures

None.

## Data Availability Statement

The data supporting the findings of this study were provided by the Fire and Disaster Management Agency (FDMA) of Japan. Restrictions apply: the data were used under license for the current study and cannot be redistributed by the authors. Researchers may apply directly to the FDMA; access is granted at the FDMA’s discretion through its own formal application and approval process, independent of the study authors, and may require a signed data access agreement.

## Notes

### Competing Interest Statement

The authors have declared no competing interest.

### Author Declarations

This study was approved by the Institutional Review Board of Meiji University of Integrative Medicine (No. 2026-030). The data supporting the findings of this study were provided by the Fire and Disaster Management Agency (FDMA) of Japan. Restrictions apply: the data were used under license for the current study and cannot be redistributed by the authors. Researchers may apply directly to the FDMA; access is granted at the FDMA's discretion through its own formal application and approval process, independent of the study authors, and may require a signed data access agreement.

